# Impact of weight-bearing and sex-stratified differences in risk factors of bone loss on bone mineral density in HIV conditions – findings of the Nigeria HIV-BMD study: An observational study

**DOI:** 10.1101/2022.10.13.22281045

**Authors:** Sam Chidi Ibeneme, Gerhard Fortwengel, Ifeoma Joy Okoye, Wilfred Okwudili Okenwa, Amarachi Destiny Ezuma, Akachukwu Omumuagwula Nwosu, Georgian Chiaka Ibeneme, Amaka Nnamani, Dnyanesh Limaye, Firas Fneish, Hellen Myezwa, Okere Philip, Nneka Iloanusi, Adedayo Tunde Ajidahun, Ifeoma Ulasi

## Abstract

**Background:** Biomechanical loading exerts an osteogenic stimulus; thus, bone mineral density(BMD) may vary in weight-bearing and non-weight-bearing bones. Therefore, weight-bearing activities could modulate sex-, HAART- and HIV-related BMD loss.

**Method:** A cross-sectional observational study of 503 people living with HIV (PLWH) selected by convenience sampling at Enugu State University Teaching Hospital, Nigeria, was conducted from September 2015 to September 2016. The BMD of toe or weight-bearing(BMD_toe_) and thumb or non-weight-bearing(BMD_thumb_) bones were measured with Xrite 331C densitometer and compared using independent t-test. Impact of the risk factors (age, weight, body mass index-BMI, duration of HIV, height and types of HAART) of bone loss and their relationships with the BMD were compared across the sexes using multivariate, and univariate regression analyses, at p<0.05,two-tailed.

**Result:** Participants comprised of females(378/75.1%), males(89/17.7%) and others(36/7.16%) without gender specificity, with mean age=37.2±9.79years, and BMI=25.6±5.06kg/m. HAART-experienced participants’ (352/69.98%) mean HAART-exposure duration was 4.54±3.51years. BMD_toe_(−0.16±0.65g/cm^3^) was higher(p<0.05) than BMD_thumb_(−0.93±0.44g/cm^3^), and differed across the BMI classes (p=0.000003;d=0.998), and was accounted for in *post hoc* analysis by normal weight versus underweight BMI classes (p=<0.001). BMD_toe_ was positively correlated with height (r=0.13,r^2^=0.0169;p<0.05), and males were taller than females(p<0.001). Females accounted for 90%(9/10) cases of osteopenia and 71.43%(5/7) osteoporosis. Males were older(p=0.002) while females had greater BMI (p=0.02), lower median BMD_toe_(p=0.005) and BMD_thumb_ (p=0.005).

**Conclusion:** Higher BMD in weight-bearing bones, and lower BMD_toe_ in underweight (sub-optimal loading) BMI class suggest a role for osteogenic stimulus and fat metabolism in bone loss. Females being younger/heavier, would have greater loading/osteogenic stimulus reinforced by lesser age-related BMD changes. Males being taller would have greater bone marrow adipose tissue that promote osteogenesis through paracrine mechanisms. Therefore, higher BMD in males should be partly explained by height-related metabolic surrogates and sex-hormonal differences. Greater BMD In females’ weight-bearing bones implies that loading ameliorates physiological tendencies towards lower BMD.

## Background

A recent World Health Organisation (WHO) global report [1] revealed that the number of people living with human immune deficiency virus-HIV (PLWH) at the end of 2018, was 37.9 million out of which 25.7 million were from Africa. Similarly, 1.1 million (two-thirds) of the 1.7 million newly infected persons were from Africa. Nigeria, the most populous African country, has 1.9 million PLWH. The World Health Organisation (WHO) also reported that in 2017, 21.7 million PLWH had access to antiretroviral therapy (ART) with Africa accounting for 15.3 million (70 %) [2,3]. The number of PLWH on ART was scaled up to 23.3 million by the end of 2018, which has so far saved 13.6 million lives from 2000 – 2018. The advent of highly active antiretroviral therapy (HAART) has significantly altered the prognosis of human immune deficiency virus (HIV) infection, enabling PLWH to live longer, but HAART has been linked to bone demineralization in several studies [4,5]. Other researchers suggested that the cause of bone loss in HIV is multifactorial including the direct effects of the virus on bone metabolism [6], which can be modulated or amplified by sex [7], age [8], types of antiretrovirals (ARVs) [9], and duration of ARV [10]. However, no study has considered that the impact of the risk factors of bone loss may vary in weight-bearing and non-weight-bearing bones, which may warrant specific intervention in each context hence this study.

HIV infection directly alters bone metabolism (and by extension the bone structure) through its immune-depressive effects, and by stimulating the release of inflammatory mediators such as cytokines, resulting in reduced bone mineral density (BMD) and bone mass [11, 12, 13-18, 19-22]. This action in the absence of an opposing anti-inflammatory response will distort bone homeostasis [23]. Increased cytokine levels in HIV infection, particularly the pro-inflammatory cytokines – interleukin-1 (IL-1), interleukin-6 (IL-6), tumour necrosis factor-alpha (TNF-α) and RANKL (Receptor activator of nuclear factor kappa-B ligand) [24] provide evidence in support of this hypothesis. However, conclusive evidence of the role of these pro-inflammatory mediators in bone mineral loss in HIV infection is still lacking.

Certain types of antiretroviral therapy (ARVs) have direct effects on the bone, including protease inhibitors, nucleoside/nucleotide reverse transcriptase inhibitors and non-nucleoside reverse transcriptase inhibitors [11, 25-30]. However, HAART was also found to not affect the BMD by other studies [31-33]. The duration of HAART usage was linked to the stability of the BMD over time. The literature abounds with conflicting reports on this issue [34,35]. A study [33] claims that the BMD initially decreased within the first two years of commencing HAART but increased and stabilized thereafter. The authors [34] also reported that changes in body weight were related to greater changes in BMD. Other authors [35] claimed that no change in the BMD occurs till about two and half years after the commencement of HAART, and thereafter instability sets. The authors [35] further reported that low body weight accounted for the low BMD. The disparity in the studies [34,35] was attributed to HAART-related weight gain, which in longitudinal studies would either report stable or increasing BMD, and likewise in cross-sectional studies conducted after an elongated time interval following the commencement of HAART. In contrast, the BMD will be reported as low in cross-sectional studies conducted within a short time after HAART has commenced, in which period there was little or no HAART-related weight gain. [35]. The credence of this view was buttressed by the fact that the only study [36] on an HIV-infected group with a comparable weight-matched control group showed no difference in the BMD between the groups. Therefore, weight and body mass index are risk factors for low BMD in PLWH [37, 32, 38].

The association of age with BMD is in the literature consistent, irrespective of whether the prevalence was high or low. For instance, The CDC-funded SUN Study revealed that osteoporosis was associated with older age in a US population cohort of PLWH in the HAART era [39]. Another study [40] that examined BMD and incident fractures in a cohort of 328 PLWH and 231 healthy controls, not only found significantly lower BMD in the PLWH, but that age was an important predictor of low BMD in this Population. The two studies above demonstrate that traditional risk factors would have a more significant impact on the BMD of PLWH than ART-related factors [41]. Gender is also a traditional risk factor for low BMD and the female gender is an independent predictor of low BMD in PLWH [42]. Similarly, the female gender is also an independent predictor of hypovitaminosis D, while low BMD is associated with comorbidity hypovitaminosis D [43]. Therefore, the physiological attributes of the female could be the key driver of the relationship with BMD in PLWH.

A key remedy to bone loss is weight-bearing activities which refer to physical activities that make an individual move against gravity while staying upright with the feet on a supporting surface. Weight-bearing activities (standing) significantly increased BMD in lower extremities of patients with spinal cord injury [44], stroke patients [45], athletes [46], and older women [47] and thus represent a powerful osteogenic stimulus that could have translational benefit in PLWH. Consequently, the impact of the risk factors of low BMD in PLWH should vary in weight-bearing and non-weight-bearing bones. Such variations could be of sufficient significance to warrant specific interventions in PLWH. Therefore, we sought to i. Assess the variations in the BMD of weight-bearing (BMD_toe_) and non-weight-bearing bones (BMD_thumb_) in PLWH, ii. Assess the impact of some risk factors of bone demineralization (sex, age, BMI, HAART types, and duration of HAART) on BMD of weight-bearing and non-weight-bearing bones, iii. Determine the sex-stratified differences in BMD in relation to some risk factors of bone loss (age, height, weight, BMI, drug duration, type of drug and duration of HIV) in weight-bearing and non-weight-bearing bones..

## Methods

### Study design, study population and sample size

This is a cross-sectional observational study assessing bone mineral density in PLWHA (who were HAART-experienced and HAART-naive). The study is the first phase of the Nigeria HIV-BMD study, which started in September 2015 to September 2016. It was based primarily at the Enugu State University Teaching Hospital, Parklane, Enugu but participants were recruited from other clinics within the State offering HIV services. Enugu State has an estimated population of 3.8 million and is one of the 36 states of Nigeria. It is located in the southeastern part of Nigeria with a predominantly Igbo-speaking population. The State has two university teaching hospitals - the University of Nigeria Teaching Hospital (UNTH) Enugu and the Enugu State University Teaching Hospital and College of Medicine. The State also has seven District Hospitals and at least one health centre or cottage hospital in each of the 17 Local Government Areas, in addition to several private hospitals and clinics. At the time of this study, Enugu State had an HIV prevalence of 1.3% (Male: 1.0%; Female: 1.6%) [48] with about 34,268 people living with HIV [48]. About 20,297 people are living with HIV on treatment in 21 treatment facilities. The total number of facilities offering HIV services in the State is 318. Recent data showed that Enugu State’s HIV prevalence is 2.1% with about 66,768 PLWH [49].

The study population consisted of all persons (HIV positive and HIV negative) aged 18 – 60 years attending the general outpatient clinic or the HIV clinic of the University of Nigeria Teaching Hospital Enugu, or the outpatient clinic or HIV clinic of one of the health facilities in and around Enugu. For the HIV-positive participants, a documented positive HIV result plus a history of being followed up at the HIV treatment and care centre was sufficient criteria for enrolment into the study and classification as HIV positive, provided other inclusion criteria are met. For newly diagnosed HIV patients without a history of clinic follow-up, a repeat HIV test was done to confirm the diagnosis of HIV.

#### Eligibility Criteria

##### Inclusion Criteria

The inclusion criteria were: i. At least 18 years at the last birthday and not more than 60 years at the next birthday regardless of gender; ii. PLWH who are HAART-experienced.

##### Exclusion Criteria

the exclusion criteria included – i. Severely ill persons, ii. Refusal of consent, iii. Ongoing or recent treatment with corticosteroids, hormones, immunomodulators, cytotoxic agents, or diuretics within the last 6 months or other treatment that could affect or induce bone disease and iv. Presence of severe chronic illnesses such as chronic renal failure, v. Calcium supplementation, and vi. Presence of any bone or rheumatic disorder

The sample size calculation was performed on the assumption that the prevalence rate of HIV in Enugu State is 1.3 % of the total State population [48]. At the time of this study, the actual number of people living with HIV from program data was 34,268 [50]. The sample size was calculated to derive a 95% confidence interval with a margin of error no greater than 5%. The minimum recommended sample size was determined as 760. Subsequently, consecutive patients identified from the participating clinics were approached for consent to participate in the study.

### Data collection

Once the participants’ informed consent was provided, a study nurse/physiotherapist/physician administered a screening questionnaire to determine eligibility for the study. Only those who met the inclusion criteria were recruited and required to provide their demographic data (age, sex, occupation, educational status, marital status), previous medical and surgical history (HIV status and duration, ART status & duration, history of fractures and hospitalization, chronic illnesses, ongoing and recent medications particularly corticosteroids). Where a participant reported a history of fracture, the site, date and circumstances of the fracture were requested. A clinical evaluation was then conducted on the participant by the study clinician which included a general physical examination and measurement of the weight, and height. Any evidence of opportunistic infection and lipodystrophy was documented. Once the participant has provided the demographic information, they were referred for BMD measurement using the Xrite 331C densitometer (USA).

#### i. Bone Mineral Density

BMD was measured at the right and left thumbs and big toes using an Xrite 350 densitometer. All measurements were performed with the same densitometer and by the same technician. The measurements were adjusted for age and sex and expressed as T-scores corresponding to the mean BMD for normal persons of similar age and sex minus the patient’s BMD, divided by the standard deviation (SD) of the mean for the reference population.

### Data Management and statistics analysis

Participants’ physical measurements, as well as clinical and BMD measurements, were entered into the case report form (CRF) and forwarded to the data entry clerk for entry into the Electronic Data Capture System (EDC). Participants’ data were stored on a SecurTrial server located in Berlin, Germany. The original hard copies of the questionnaires, CRFs and related documents were stored securely at UNTH Enugu in lockable filing cabinets with restricted access, both during and after the completion of the study. All the study documents were stored for a minimum of five years after the publication of the primary data.

Using the R statistical software 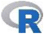, BMD in HAART-experienced PLWH was compared, controlling for age and sex. Means and standard deviations were used to express summary statistics for normally distributed continuous variables, while geometric means and standard error of mean were used to summarize skewed continuous variables after transformation. Where transformation did not correct the skewness of the data, the summary statistics were reported as medians and interquartile range (IQR). Test of significance for discrete data were carried out using the chi-square test. The Chi-square for the contingency table was employed to determine the level of independence of the independent variables. Independent t-tests were used to compare quantitative data across groups. The impact of HIV disease on BMD was tested using multivariate analysis after adjusting for demographic, anthropometric, treatment-related, HIV-related and pathophysiologically plausible variables. Only variables attaining a p-value of <0.2 in univariate analysis were included in the final multivariate model. To assess the impact of HAART treatment on BMD, the participants’ data were stratified based on the type of treatment being given. In the first instance, the HAART-experienced PLWH were categorized according to the type of drug types they received including: Nucleoside Reverse Transcriptase Inhibitors (NRTI); Non-Nucleoside Reverse Transcriptase Inhibitors (NNRTI); Protease Inhibitors (PI) and then compared in a regression analysis. The impact of the durations of HAART exposure on BMD was determined and compared in weight bearing and non-weight bearing bones using regression analysis. The relationship between the T-score and risk factors of low BMD (age, height, gender, BMI, drug duration and drug type) was evaluated, respectively using univariate regression analysis. All tests were two-sided, with a significance level of 0.05.

### Ethical consideration

This study was approved by the University of Nigeria Health Research Ethics Committee on certificate number – NHREC/05/01/2008B. Participants’ confidentiality was maintained by using code numbers instead of names, and ensuring that records were destroyed at the end of the study. Studies reviewed included only those where the participants gave their written informed consent, before participation, and after the purpose of the study was explained to them. They were informed of their right to withdraw from the study at any time of their choice, and these rights were strictly respected following the Helsinki declaration.

## RESULTS

Participants’ flow through the study is presented in Figure 1. The study participants were 503 comprising 378 females (75.10 %), 89 males (17.70 %), and 36 (7.16 %) without gender specificity. The participants’ flow through the study is presented in Figure 1. The proportion of the females was significantly higher (p < 0.0002) than males and the ratio of males to females was 1:4. The participants’ demographic characteristics were presented in Table 1. Missing data were observed as some participants did not provide all the required information, and their wishes were respected. Participants on HAART were 352 (out of 503 participants) including those on NNRTI (n= 10 or 2.84 %), NRTI (n = 44 or 12.5 %, PI (n = 31 or 8.81 %), and NRTI+NNRTI (n = 267 or 75.9 %). A significant proportion (p < 0.0002) of them were on NRTI+NNRTI.

**Fig. 1:**
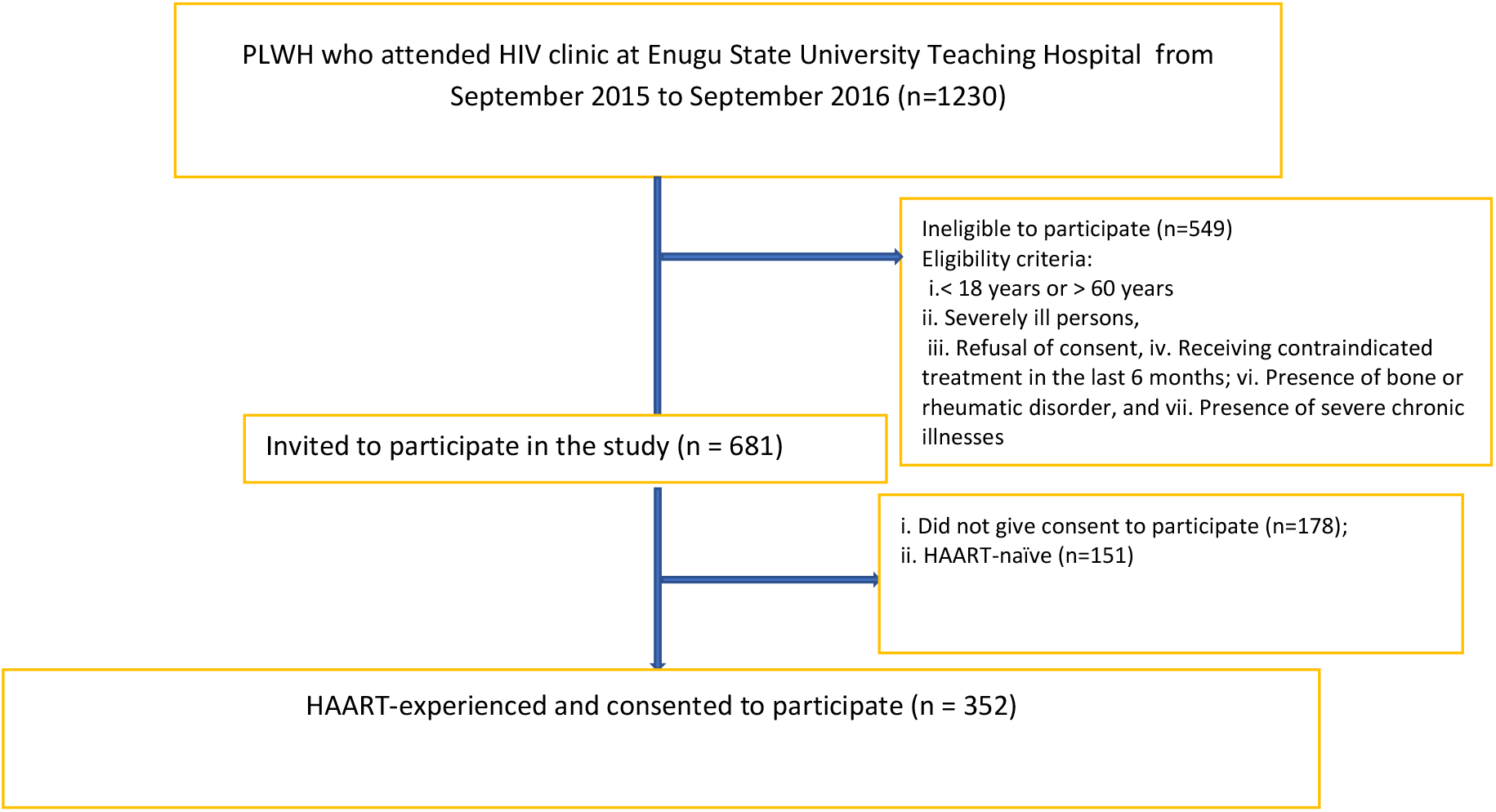
Design and flow of participants through the study

**Table 1:** Physical and clinical characterization of all participants (**N=503)**

| Variables | Number (%) | Mean (SD) | Z-value | 95% CI | p |
| --- | --- | --- | --- | --- | --- |
| Sex |  |  |  |  |  |
| Male | 89 [17.7%] |  |  |  |  |
| Female | 378 [75.1%] |  | 15.95 | 0.44 – 0.55 | < 0.0002*** |
| Unknown | 36 [7.16%] |  |  |  |  |
| **Total** | **503** |  |  |  |  |
| Age (years) | 446 | 37.2 [9.79] |  |  |  |
| Weight (kg) | 475 | 65.9 [14.3] |  |  |  |
| Height (m<sup>2</sup>) | 475 | 1.60 [0.09] |  |  |  |
| BMI (kg/m<sup>2</sup>) | 475 | 25.6 [5.06] |  |  |  |
| Duration (years) | 256 | 4.54 [3.51] |  |  |  |
| Drugs type |  |  |  |  |  |
| NRTI | 44 [12.5%] |  |  |  |  |
| NRTI+NNRTI | 267 [75.9%] |  | 13.719 | 0.45 – 0.58 | < 0.0002*** |
| NNRTI | 10 [2.84%] |  |  |  |  |
| PI | 31 [8.81%] |  |  |  |  |
| **Total** | **352** |  |  |  |  |
| BMD T-score |  |  |  |  |  |
| Thumb | 458 | -0.93 [0.44] |  | 0.70 - 0.84 | <.000001***** |
| Toe | 458 | -0.16 [0.65] |  |  |  |

**BMI:** Body Mass Index; **BMD:** Bone Mineral Density; **NRTI:** Nucleoside Reverse Transcriptase Inhibitors; **NNRTI:** Non-Nucleoside Reverse Transcriptase Inhibitors; **PI:** Protease Inhibitors.
\*\*\*signifies $p < 0.0001$ ; \*\*\*\*\*signifies $p < 0.000001$

Participants’ physical characteristics included the following: mean age = 37.2±9.79 years, and age range =10 – 60 years. Participants’ mean duration of HAART exposure = 4.54±3.51 years; and the range was 0 - 16 years, and thus, most participants were on HAART for about 5 years.

The mean age of the participants was 37.2±9.79 years with an age range of 10–60 years. Hence, the patients were relatively young adults and of productive age. In our study, a total of 352 participants reported having been placed on HAART (Highly Active Antiretroviral Therapy) with the significant majority (p < 0.0002) of them on combined therapy (NNRTI and NRTI). The mean duration of HAART exposure was 4.54±3.51 years with a duration range of 0 to 16years. Thus, most participants were on HAART for about 5 years. The mean BMI was 25.6±5.06 kg/m^2^. Furthermore, the mean BMD_thumb_ (T-score) was -0.93±0.44 g/cm_3_ and significantly (p < 0.000001) differed from the BMD_toe_ (−0.16±0.65 g/cm^3^), respectively. The scores illustrate a lower BMD at the thumbs than at the toes.

### Distribution of the BMD across risk factors in weight-bearing and non-weight bearing bones

Out of four hundred and fifty-eight participants tested for their BMD, 437/475 (92%) presented with a normal BMD_toe_ t-score while 19/475 (4%) were classified as having either osteopenia or as osteoporosis (Table 2). The BMD values suggest that cases of osteopenia and osteoporosis involved only the thumb (i.e., non-weight bearing bones) while the BMD of the toes (i.e., weight bearing bones) were within normal range (−1 to + 1). The mean BMD_toe_ t-score value was – 0.16±0.65 and was not normally distributed (Shapiro-Wilk test for normality, p = -0.00057) (Figure 1). Similarly, the mean BMD_thumb_ was -0.93±0.44 (Table 1), and was not normally distributed, (Shapiro-Wilk Test for normality, p= -0.084) (Figure 1). The cases of osteoporosis and osteopenia varied across several demographic characteristics (Table 2). The females accounted for nine (90%) out of the ten cases of osteopenia, and five (71.43%) out of seven cases of osteoporosis compared to the males.

**Table 2.** Differences in BMD of weight-bearing and non-weight bearing bones across some risk factors of bone loss

| Variables | No of PLWH | No with Normal BMD | No with Osteopenia | No with Osteoporosis | Mean BMD<sub>thumb</sub> (g/cm<sup>2</sup>) | 95% CI | BMD<sub>thumb</sub><br>F | P-value | Mean BMD<sub>toe</sub> (g/cm<sup>2</sup>) | BMD<sub>toe</sub><br>95% CI | F | p |
| --- | --- | --- | --- | --- | --- | --- | --- | --- | --- | --- | --- | --- |
| **Sex** |  |  |  |  |  |  |  |  |  |  |  |  |
| Female | 378 | 349 | 9 | 5 | -0.98 | Ref |  | Ref | -0.18 | Ref |  | Ref |
| Male | 89 | 82 | 1 | 2 | -0.76 | 0.11- 0.31 |  | <0.001*** | -0.06 | -0.03 - 0.28 |  | 0.112 |
| **Total** | **467** | **431** | **10** | **7** |  |  |  |  |  |  |  |  |
| **Age Group** |  |  |  |  |  |  | 1.08 | 0.34 |  |  | 0.70 | 0.49 |
|  |  |  |  |  |  |  |  | **Post-hoc** |  |  |  | **Post-hoc** |
| (<21 years) | 10 | 10 | 0 | 0 | -1.00 | Ref |  | Ref | -0.23 | Ref |  | Ref |
| (21-40 years) | 286 | 266 | 3 | 7 | -0.96 | -0.21-0.35 |  | 0.616 | -0.13 | -0.32 - 0.51 |  | 0.660 |
| (>40 years) | 150 | 138 | 7 | 0 | -0.90 | -0.15-0.42 |  | 0.365 | -0.21 | -0.40 - 0.44 |  | 0.940 |
| **Total** | **446** | **414** | **10** | **7** |  |  |  |  |  |  |  |  |
| **BMI Classes** |  |  |  |  |  |  | 1.51 | 0.19 |  |  | 6.64 | 3.371e-05*** |
|  |  |  |  |  |  |  |  | **Post-hoc** |  |  |  | **Post-hoc** |
| Underweight | 18 | 13 | 1 | 3 | -1.10 | -0.45-0.01 |  | 0.064 | -0.60 | 0.47 - 1.10 |  | <0.001*** |
| Normal | 237 | 217 | 7 | 3 | -0.89 | Ref |  | Ref | -0.18 | Ref |  | Ref |
| Overweight | 126 | 117 | 3 | 1 | -0.98 | -0.19-0.01 |  | 0.067 | -0.23 | -0.18 - 0.10 |  | 0.547 |
| Obesity I | 90 | 86 | 1 | 0 | -0.92 | -0.13-0.09 |  | 0.690 | -0.16 | -0.13 - 0.18 |  | 0.785 |
| Obesity II | 4 | 4 | 0 | 0 | -0.97 | -0.52-0.36 |  | 0.731 | -0.09 | -0.53 - 0.72 |  | 0.756 |
| **Total** | **475** | **437** | **12** | **7** |  |  |  |  |  |  |  |  |
| **HAART Duration** |  |  |  |  |  |  | 0.3 | 0.81 |  |  | 0.53 | 0.66 |
|  |  |  |  |  |  |  |  | **Post-hoc** |  |  |  | **Post-hoc** |
| (0-4 years) | 146 | 143 | 3 | 0 | -1.04 | -0.06-0.12 |  | 0.474 | -0.29 | -0.12 - 0.07 |  | 0.608 |
| (5-9 years) | 79 | 75 | 3 | 1 | -1.07 | Ref |  | Ref | -0.26 | Ref |  | Ref |
| (10-14years) | 30 | 30 | 0 | 0 | -0.94 | -0.01-0.27 |  | 0.061 | -0.30 | -0.20 - 0.10 |  | 0.516 |
| (>14 years) | 1 | 1 | 0 | 0 | -1.22 | -0.79-0.50 |  | 0.660 | -0.45 | -0.88 - 0.5 |  | 0.582 |
| **Total** | **256** | **249** | **6** | **1** |  |  |  |  |  |  |  |  |

Underweight = <18.5Kg/m<sup>2</sup>; Normal BMI = 18.5-24.9 Kg/m<sup>2</sup>; Overweight = 25.0- 29.9 kg/m<sup>2</sup>; Obesity I = 30-40 kg/m<sup>2</sup>; Obesity II = 40.1-50 kg/m<sup>2</sup>; \*indicates significance at p<0.05; \*\* indicates significance at p<0.01; \*\*\* indicates significance at p<0.001, **NB**. The highlighted categories indicate references for each of the variables.; No with Osteoporosis = > - 2.5 g/cm<sup>2</sup>; No with Osteopenia = 1.0 to > - 2.5 g/cm<sup>2</sup>; Normal BMD = +1 to -1 g/cm<sup>2</sup>

### BMD and age

The age distribution of bone demineralization based on the BMD_thumb_ showed that all cases of osteoporosis (7/100%) occurred in participants aged 21 – 40 years, while 70% (7/10) of those with osteopenia were > 40 years old (the policy at that time is that newly infected PLWH are not placed on HAART until viral load of 200 copies/ml is reached). However, the mean BMD_thumb_ (−1.03) of those aged < 21 years old suggested the prevalence of osteopenia. Statistical analysis of the data showed that BMD_toe_ and BMD_thumb_ did not differ significantly across age groups.

### BMD and BMI classes

We explored the BMD distribution across the BMI classes after categorizing the participants according to WHO BMI classification: underweight = <18.5 kg/m^2^; normal weight =18.5-24.9 kg/m^2^; overweight =25.0-29.9 kg/m^2^; obese I =30-40 kg/m^2^ and obese II =40.1-50 kg/m^2^ (Table 2). The BMI distribution among the 475 participants was as follows: normal weight (n = 237 or 49.9 %), underweight (n = 18 or 3.79%), overweight (n=126 or 26.5%), obese class I (n = 90 or 18.9%), and obese class II (n=4 or 0.84 %) (Table 1). The mean BMI = 25.6±5.06 kg/m^2^. Furthermore, we found no significant differences when we explored the impact of the duration of HAART exposure on the BMI (F (4, 250) = 1.64; p = 0.16). BMD distribution across the BMI classes showed that the highest (3 out of 7 or 42.86%) case of osteoporosis was reported among some underweight and normal weight participants, respectively. However, it was only the mean BMD_thumb_ (−1.10) of the underweight participants that suggested the prevalence of osteopenia compared to other BMI classes.

The one-way ANOVA test (Table 2) showed a significant variation in the mean BMD_toe_ across the BMI classes (F (4, 451) = 6.6398; p = 0.000003.371), respectively. The *post-hoc* analysis showed a significant mean difference in the BMD_toe_ for normal weight versus underweight BMI classes (p = < 0.001), overweight versus underweight, and obese class I versus underweight (Fig. 2a). The effect size was large for normal versus underweight classes (d = 0.998), overweight versus underweight classes (d = 0.964), and obese class I versus underweight classes (d = -0.796). In contrast, the BMD_thumb_ did not vary across the BMI classes (Fig, 2b).

**Figure 2a.**
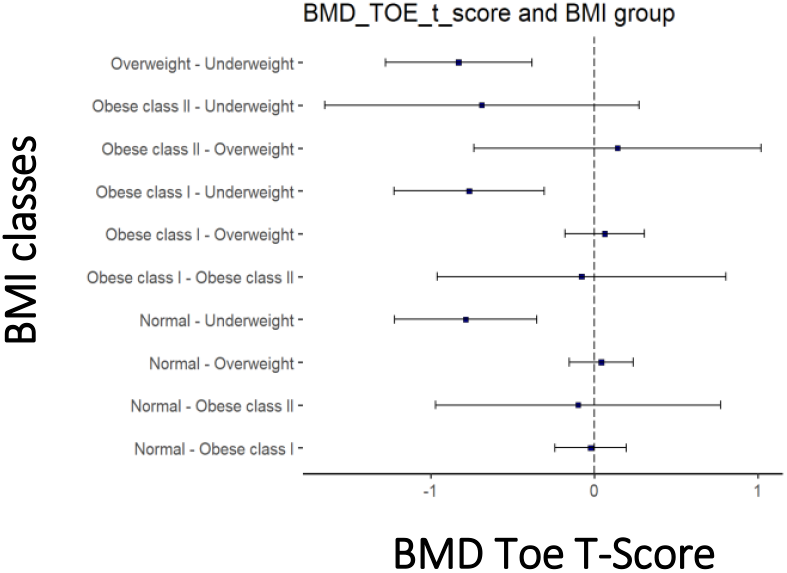
Whisker plot to show the mean differences and relationship between BMI classes and toe BMD T-score.

**Figure 2.**
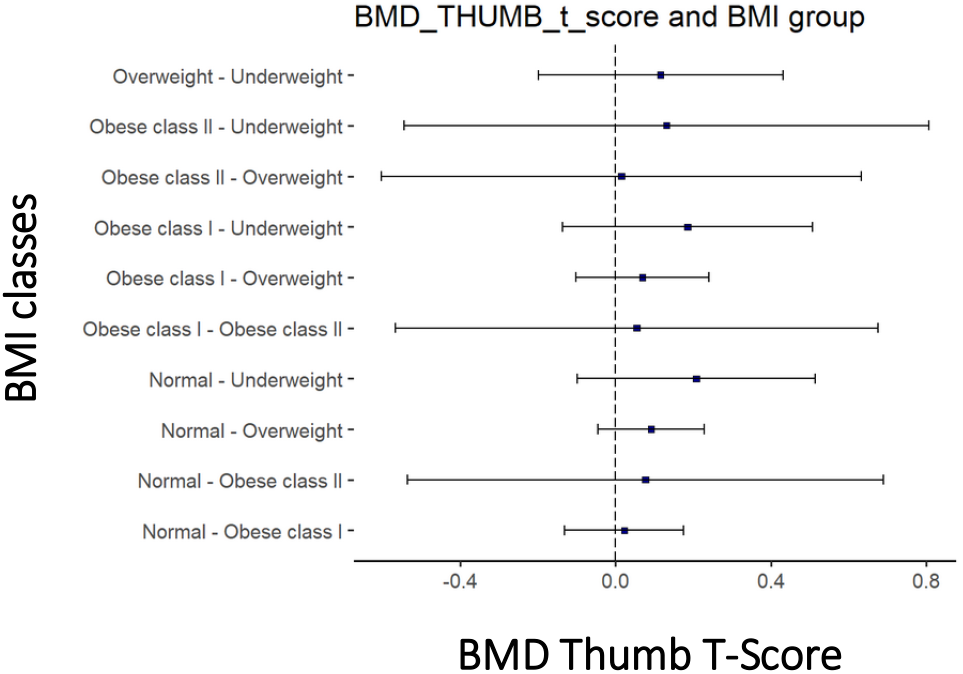
Whisker plot to show the mean differences and relationship between BMI classes and Thumb BMD T-score.

### BMD and duration of HAART

The BMD distribution across the duration of HAART exposure was evaluated and showed some cases of osteoporosis among participants with 5 – 9 years duration of HAART exposure. Participants mean duration of HAART exposure = 4.54±3.51 years; and range= 0 - 16 years. The highest cases of osteopenia (50% or 3 out of 6) were found in those with 0 – 4 years, and 5 – 9 years of HAART exposure, respectively. However, some cases of osteopenia for BMD_thumb_ were found in participants exposed to HARRT for 0 – 4 years (−1.04), 5 – 9 years (−1.07) and > 14 years (−1.22). The BMD did not differ significantly across the categories of the duration of HAART exposure (F (3,252) = 0.22; p=0.88) as shown in Figures 3a and 3b.

**Figure 3a.**
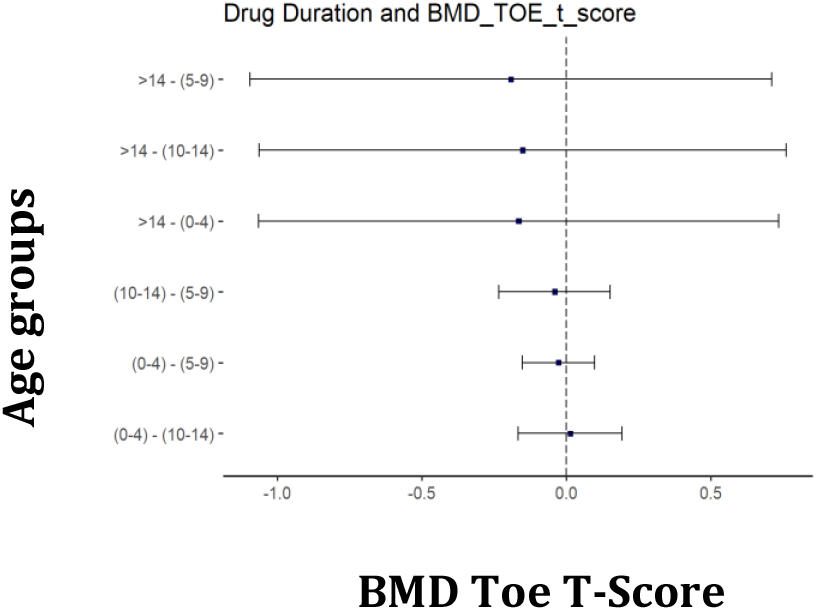
Whisker plot to show the mean differences and relationship between drug duration and toe BMD T-score.

**Figure 3b.**
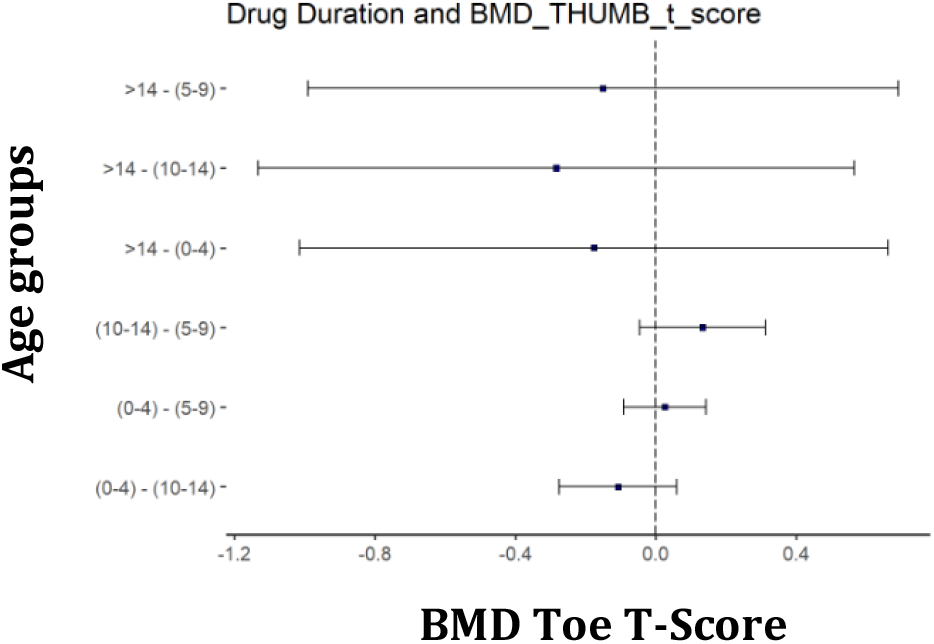
Whisker plot to show the mean differences and relationship between drug duration and thumb BMD T-score.

### BMD and classes of HAART

A further analysis (Fig. 4a and 4b) of the impact of the classes or types of HAART taken on the BMD_toe_ (Fig. 4a) showed no significant difference (F = 0.53; p = 0.66) similar to the BMD_thumb_ (F = 0.33; p = 0.66) (Fig. 4b).

**Figure 4a.**
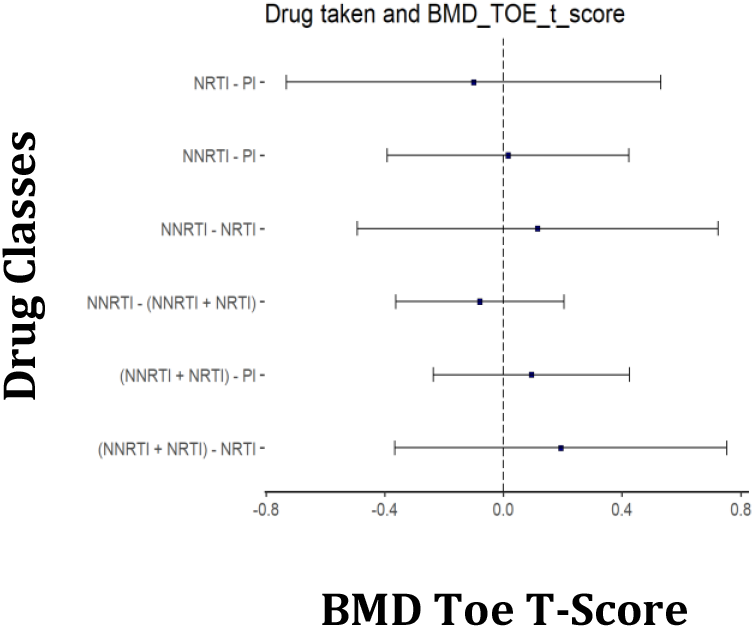
Whisker plot showing the mean difference between HAART and toe BMD T-score.

**Figure 4b.**
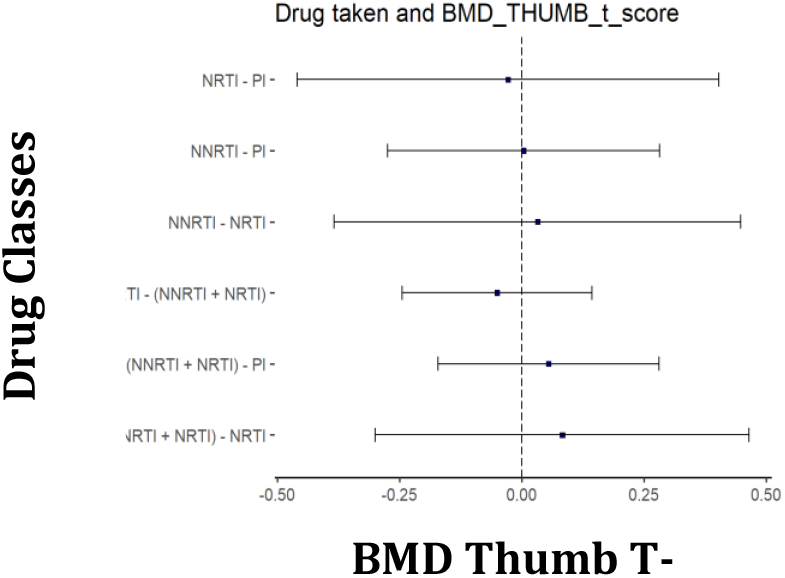
Whisker plot showing the mean difference between HAART and thumb BMD T-score.

### Relationship between the BMD and some risk factors of bone loss

**T**he overall univariate analysis (Fig. 5) without regard to gender was conducted. The BMD_toe_ was significantly and positively correlated with height (r = 0.13, P < 0.05). Duration of exposure to HAART was significantly and positively related to BMI (r = 0.14, P < 0.05), weight (r = 0.15, P < 0.05), and age (r = 0.27; p < 0.0001). BMI was positively related to age (r = 0.15; p < 0.05) and weight (r = 0.88; p = < 0.0005). Age was positively related to weight (r = 0.18; p = < 0.005), but was negatively and insignificantly related to BMD_toe_ (r = - 0.023; p > 0.05). The univariate analysis further showed a negative but insignificant correlation between the BMD_thumb_ and BMI (r = - 0.084; p > 0.05) as well as BMD_thumb_ and weight (r = - 0.061; p > 0.05) among PLWH.

**Figure 5.**
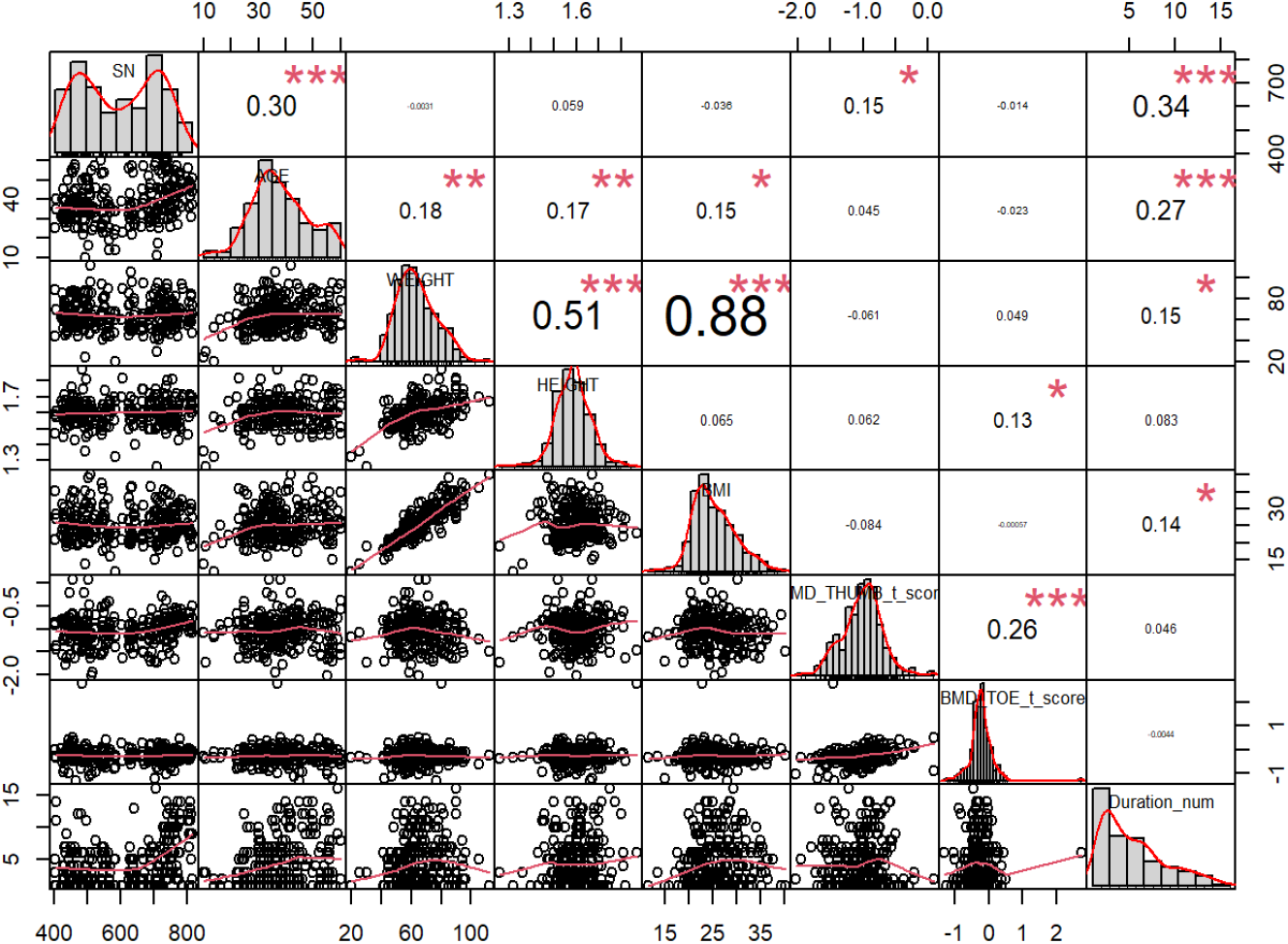
Correlation between variables for PLWHIV.

### Sex-stratified relationship between the BMD and some risk factors of bone loss Females

In the females, the univariate analysis showed that the BMDtoe was negatively related to the duration of HAART, age, and height while the BMD_thumb_ was negatively related to age, height, weight and BMI but positively related to the duration of HAART Table 3). However, the differences were not significant (p > 0.05). Similarly, the BMI was positively related to the duration of HAART but the relationship was also insignificant (p > 0.05).

**Table 3:** Relationship between the BMD and some risk factors of bone loss in females

| Variables | Age | Weight | Height | BMI | BMD<sub>thumb</sub> | BMD<sub>toe</sub> | Duration |
| --- | --- | --- | --- | --- | --- | --- | --- |
| Age |  |  |  |  |  |  |  |
| Weight | 0.15* |  |  |  |  |  |  |
| Height | 0.023 | 0.47*** |  |  |  |  |  |
| BMI | 0.17 | 0.92*** | 0.09 |  |  |  |  |
| BMD<sub>thumb</sub> | -0.046 | -0.13<sup>#</sup> | -0.081 | -0.098 |  |  |  |
| BMD<sub>toe</sub> | -0.065 | 0.019 | -0.026 | 0.039 | 0.30*** |  |  |
| Duration | 0.24*** | 0.12<sup>#</sup> | 0.035 | 0.12<sup>#</sup> | 0.065 | -0.048 |  |

\*indicates significance at $p < 0.05$ ; \*\* indicates significance at $p < 0.001$ ; \*\*\* indicates significance at $p < 0.0001$

### Males

In the males (Table 4), the univariate analysis showed that the BMD_toe_ was negatively correlated with age and positively related to height, weight and BMI, and duration of HAART though the relationship was not significant (p > 0.05). The BMD_thumb_ was positively related to age, weight, height and BMI but negatively correlated to the duration of HAART. Nevertheless, the relationship was not significant. However, a significant positive relationship was established between BMI and duration of HAART (r = 0.28, p = < 0.05), but the proportion of variance was small (r^2^ = 0.08).

**Table 4:** Relationship between the BMD and some risk factors of bone loss in males

| Variables | Age | Weight | Height | BMI | BMD<sub>thumb</sub> | BMD<sub>toe</sub> | Duration |
| --- | --- | --- | --- | --- | --- | --- | --- |
| Age |  |  |  |  |  |  |  |
| Weight | 0.27* |  |  |  |  |  |  |
| Height | 0.31 | 0.72*** |  |  |  |  |  |
| BMI | 0.23<sup>#</sup> | 0.85*** | 0.27<sup>#</sup> |  |  |  |  |
| BMD<sub>thumb</sub> | 0.19 | 0.13 | 0.14 | 0.14 |  |  |  |
| BMD<sub>toe</sub> | -0.022 | 0.11 | 0.20 | 0.0078 | 0.15 |  |  |
| Duration | 0.32* | 0.27 | 0.14 | 0.28* | -0.047 | 0.051 |  |

\*indicates significance at $p < 0.05$ ; \*\* indicates significance at $p < 0.001$ ; \*\*\* indicates significance at $p < 0.0001$

### Sex-stratified distribution of BMD and some risk factors for bone loss

We evaluated the sex-stratified variations in some risk factors of low BMD and found (Table 5) that the median age for the males (42.0 (IQR, 34.0-49.0) differed significantly (p = 0.002) from the females (35.0 (IQR, 30.0-42.0) similar to median height [(females = 1.60 metres (IQR, 1.55; 1.63); males = 1.69 metres (IQR, 1.62; 1.73); p = < 0.001) (Table 5). The female median BMI (24.8 kg/m^2^ (IQR, 22.1-28.8), differed significantly (p = 0.02) from the males (23.2 kg/m^2^ (IQR, 21.2-26.8) (Table 3) unlike the median weight (p= 0.08). The male median duration of HAART exposure [4.00 (IQR, 1.00; 6.00) years], did not differ (p=0.356) from the females [3.00 (2.00; 6.00) years]. The median BMD_toe_ T-scores for females [-0.24 g/cm^3^ (IQR, -0.42- (−0.02)] was significantly lower (p = 0.005) compared to males [-0.13 g/cm^3^ (IQR, - 0.35 - 0.18)], Similarly, the median BMD_thumb_ T-scores for females [-1.00 g/cm^3^ (IQR, -1.24; - (−0.78)] was also significantly lower (p = 0.005) than males [--0.83 g/cm^3^ (IQR, -1.05 – (−0.58)].

**Table 5.**
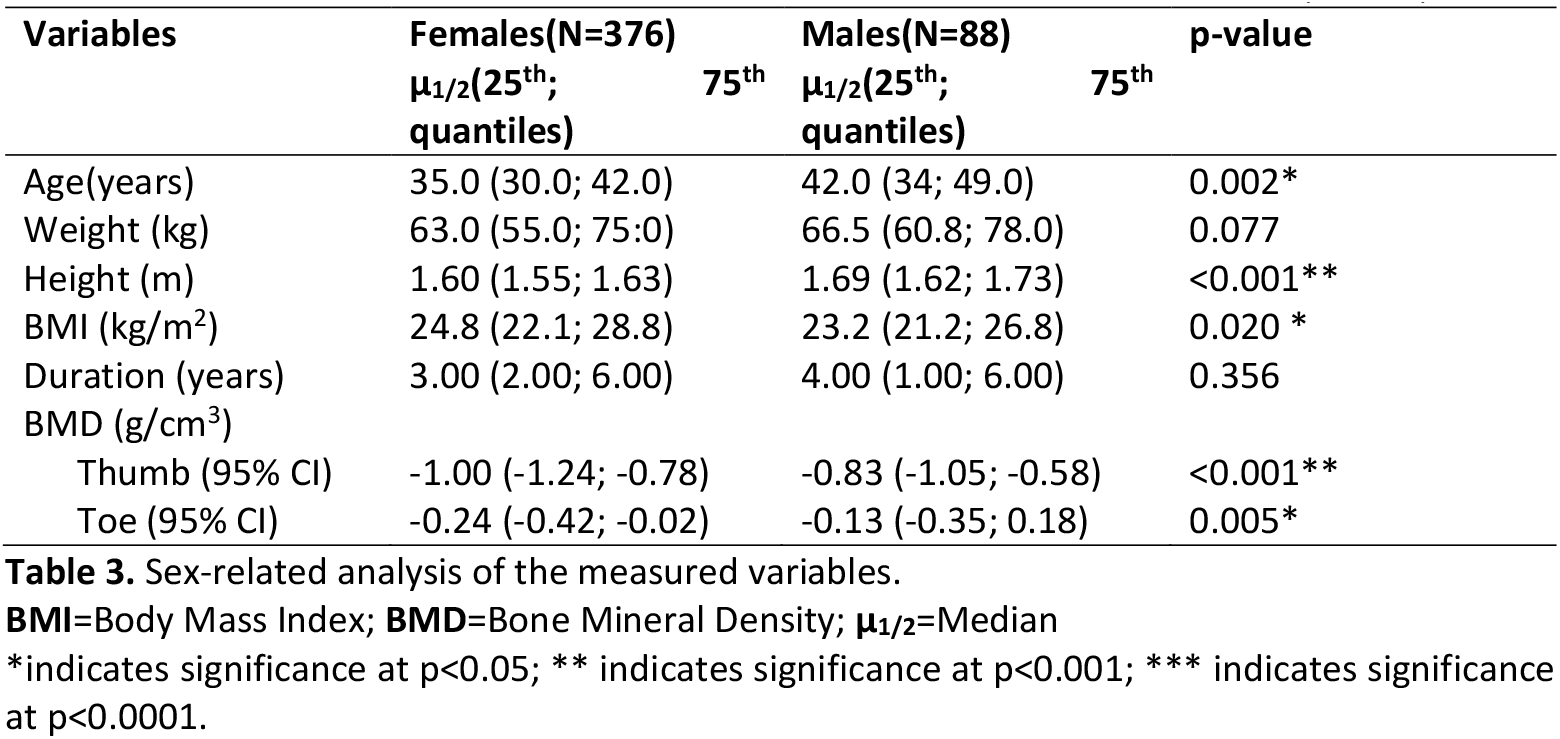
sex-stratified differences in the BMD and some risk factors of bone loss (**N=503**)

## DISCUSSION

This study mainly sought to determine the variations of the BMD in weight-bearing and non-weight-bearing bones of PLWH in relation to sex-stratified differences in the risk factors of bone loss. The main finding of this study is that the BMD of the weight-bearing bones (BMD_toes_) were significantly higher than the BMD of the non-weight-bearing bones (BMD_thumbs_). Weight-bearing activities which result in limb loading, such as walking, running, and close-chain kinetic movements [51-53], could provide a very potent osteogenic stimulus in PLWH, and is, therefore, bone protective. Consequently, weight-bearing activities could have beneficial effects on bone health in PLWH and explain why a previous study found that physical activity level is a major determinant of bone mass in postmenopausal women [54].

The physiological basis of this observation is that skeletal tissue is highly sensitive to its mechanical environment, and biomechanical strain will ensure bone mineral homeostasis [55], elicit bone formation by reducing apoptosis and enhance the multiplication and differentiation of osteoblasts and osteocytes [56]. In contrast, physical inactivity and open-kinetic movements (common with non-weight bearing bones) results in skeletal unloading and increases adipocyte differentiation while inhibiting osteoblast differentiation [57] leading to bone loss. Therefore, to prevent bone loss, non-weight-bearing bones must be loaded from time to time in HIV conditions which highlights a key role for physiotherapists in routine care of PLWH. This understanding informed our recent and ongoing clinical trials in South Africa/Nigeria on the effects of physical exercises/activity on BMD in PLWH during lower limb fracture healing [58].

Gender differences were observed as the median BMD T-score of the thumb was significantly lower (p = 0.005) in females (−1.00 g/cm^3^) compared to males (−0.83 g/cm^3^). A similar trend was observed at the toes (females = -0.24 g/cm^3^ versus males = -0.13 g/cm^3^; p = 0.005). It implies that the males had an advantage over the females with regards to bone health not minding that they were older. This could be attributed to several factors including physiological factors and sociocultural variables.

Physiologically, males should have greater bone mass than their female counterparts due to differences in skeletal muscle mass as well as hormonal make-up of which the male (testosterone) hormones have an anabolic effect on the bones [59,60]. However, a study of PLWH in Malawi [61] reported lower BMD in males than in females while another study indicated that females had lower BMD than males [7,4]. Chisati et al., [61] findings in Malawi do not necessarily mean that the female gender was not physiologically disadvantaged but could be related to the low osteogenic stimulus probably arising from the observed low daily active lifestyle activities among male Malawians than females, which should reduce skeletal muscle loading and bone mass. Similarly, the females were heavier than the males considering their BMI, and which translates to increased biomechanical loading of the bones in the females than the males with proportionate osteogenic stimulus. Another study also found no significant association between gender and BMD [31] for the same reason, given that the physical activity levels between the study groups were equal as the differences were not significant. Therefore, the impact of weight-bearing physical activities seemed to override or modulate the physiological tendencies of the female gender towards low BMD.

The low BMD among the females studied by Erlando et al., [7] was linked to the negative effects of lack of estrogen on bone mass [5,59] as 24% of the female participants were in menopause. In essence, different reasons accounted for the diverse findings observed in the literature. Invariably, the relationship between gender and BMD in HIV conditions can be influenced by several confounders which can be manipulated to increase BMD or considered when interpreting the findings. One of the several confounders is socio-cultural practices. This is plausible because in most African cultural communities the males are culturally assigned responsibilities for manual labour-intensive physical activities than women [62]. Hence, the men in Ethiopia men spent nearly double as much time as women in moderate/vigorous activity [63]. Similarly, men in Nigeria had significantly higher physical activity levels than women [64]. Across six sub-Saharan countries, (Burkina Faso, Ethiopia, Ghana, Nigeria, Tanzania and Uganda) males were engaged in physical exercises for up to an hour unlike females [65].

An active lifestyle should increase lean muscle mass [53] and boost physical activity which recruits the skeletal muscles and increases the loading traction on the bones thereby increasing shear strain that activates osteoblastic activities [66] in the presence of putative mediators such as nitric oxide, prostaglandins E (PGE) and (PGI) I, sclerostin, Insulin-like growth factor (IGF’s), Transforming growth factor beta (TGFβ), Receptor Activator of Nuclear Factor-kappa B-Ligand (RANK-L), and osteoprotegerin (OPG). Shear stress generated with physical exercises, involving skeletal muscle contraction, induces the production of nitric oxide [67]. Nitric oxide suppresses osteoclasts’ activity [68] and promotes osteoblastic activity [67]. Similarly, shear stress induces prostaglandin synthesis [69] which stimulates osteoblastic activity through IGFs [70], and IGFs are increased early after mechanical stimulation [71]. PGE and PGI directly inhibit osteoclastic activity [72], while they activate bone remodelling through cells of the osteoblast lineage [73]. Consequently, the well-maintained muscle in males should confer an advantage on them over the females, [66]. Therefore, gender-specific physical activity interventions may be required to improve bone health in PLWH.

Sex-stratified relationships between the risk factors of bone loss and BMD revealed some important lessons. Though the BMD_toe_ was not significantly correlated with age, HAART duration and all indices of body anthropometry, certain important trends were highlighted that differentiated the males from the females. For instance, while the BMD of the weight-bearing bones were positively affected by the duration of HAART and all the indices of body anthropometry, it was negatively affected by age. In the females, the BMD of the weight-bearing bones were negatively affected by HAART and age. This implies that age-related changes in bone health is independent of sex and could be related to microstructural weakening, arising from trabecular perforation, diminishing, loss of connectivity, cortical thinning and greater porosity [74], likely undergirded by alterations in the composition and extent of collagen cross-linking [75]. Among the PLWH, the literature cited above shows that the difference between the female Malawian and female Nigerians is that the former had higher physical activity level and BMD compared to the males [61] unlike the later [62]. Therefore, high physical activity level could be the important determinant of the BMD differences between the two populations. Notwithstanding sex-related differences, a common similarity between the males and females is that the BMD of the weight-bearing bones were significantly greater than the BMD of the non-weight-bearing bones.

Another important observation is that HAART duration had a significant positive impact on the BMI in the males but was positive and insignificant in the females. Several factors could be responsible for this observation. For instance, a high baseline BMI had independently improved 30-month T lymphocytes (CD_4+_) recovery implying that a higher BMI could determine better immune reconstitution in PLWH after commencing HAART [76]. Achieving immunologic reconstitution with time implies minimizing the negative impact of HIV on BMD as already discussed above [6, 11-22]. Since the males were on HAART for a longer period of 4 years compared to 3 years recorded in the females, they should have a better immunologic reconstitution than the females hence the consequent significant positive impact of HAART duration on their BMD.

Another probable explanation is that a higher BMI implies a higher serum level of leptin [77], and leptin not only has anabolic effects on osteoblasts and chondrocytes, but indirectly modulates bone metabolism, via multiple pathways including the hypothalamus and sympathetic nervous system, adiposity/body weight, and the expression of low brainstem hormones (e.g., pituitary) [78]. Several studies seem to buttress this view. Dutta et al., [37] reported that BMI was a predictor of osteoporosis in HIV-infected women, while Choe et al., [32] showed a significant association between low bone mass and low BMI. Similarly, Meng et al., [12] identified BMI <18.5k g/m2 as an independent risk factor for low BMD in PLWH. This suggests that there could be a link between fat metabolism and bone turnover, which was linked to leptin [79]. Even at that, there are clear differences between the sexes on how they respond to leptin hinting at possible sexual dimorphism in leptin action on BMD. In obese males, the sympathoexcitatory response to leptin is well-maintained or elevated unlike in obese females where the reproductive cycle is distorted and the sympathoexcitatory response to leptin is less robust. This could explain why BMI had a positive and significant impact on males but not significant on females. These actions are mediated by sexually dimorphic variations in Neuropeptide Y (NPY) and Pro-opiomelanocortin (POMC) entrants to the paraventricular nucleus of the hypothalamus (PVN) [80]. The consequent output on BMD should reflect the same pattern and seemed to underlie the relationship between HAART duration and BMI. Nevertheless, the fact that the BMD of the weight-bearing bones in both sexes were significantly greater than the BMD of the non-weight-bearing bones suggests that biomechanical loading of the bones is beneficial to both sexes despite their physiological or hormonal differences.

We found a significant difference in the BMD_toe_ of PLWH across the classes of BMI (F=6.6398; p=0.000003371). The significant difference across the BMI classes was accounted for by normal versus underweight. However, the impact of BMI on BMD of weight-bearing bones showed a large effect size when comparing overweight versus underweight, obese class I versus underweight; and normal versus underweight classes. It suggests that the impact of low BMI on BMD of weight-bearing bones compared to normal and other higher BMI classes, is of clinical importance, respectively. Taking reference from the underweight class, the magnitude of effect suggests that a larger number of participants in the normal weight class experienced the impact of BMI on BMD followed by overweight and obese Class I categories, respectively. However, the most protective impact of BMI against bone diseases was more evident in higher BMI classes as nobody in obesity classes I and II had osteoporosis. Similar to this trend, only 0.79 % (1 out of 126) participants in the overweight class had osteoporosis compared to 1.27 % (3 out of 237) participants in the normal weight class, and 20 % (3 out of 18) in the low/underweight class. Therefore, there is a clear trend showing that as the BMI reduces from obese class II to the underweight class, there is an increase in the proportion of participants with osteoporosis. The reverse is also true.

The finding that the low BMI category (underweight) was associated with bone demineralisation in the weight-bearing bones (toes) of PLWH follows a general trend in the literature. In the Korean population, low bone mass was significantly associated with low BMI [32]. In Indian women living with HIV, BMI was a predictor of osteoporosis [37]. In Chinese PLWH, low BMI <18.5kg/m2 was an independent risk factor for low BMD [12]. Though these findings were similar to our study, Choe et al., [32] focused their study on a single cohort (the male gender) while Meng et al., [12] involved an ART-naïve group which was not available in our study. A cross-sectional study in Uganda [81] found that a unit increase in BMI was associated with the odds of a significant reduction in low BMD. The above studies demonstrate that despite the differences in research population, location and design, the impact of low BMI adversely affects bone health. Physiologically, this trend could be attributed to a high occurrence of abnormal BMD in PLWH who are underweight [82,83]. Since high BMI has been linked to immunologic reconstitution in PLWH and improved BMD, a low BMI implies the opposite, hence another study [84] determined that HIV infection of osteoblasts could be linked to an undesirable balance of bone remodeling.

Females being younger and heavier would have greater mechanical loads and proportionate osteogenic stimulus which effect should be strengthened by less adverse age-related changes in BMD. However, taller males would have greater bone marrow adipose tissue (BAT) which promotes osteogenesis through paracrine mechanisms [85]. The implications of the results is that greater mechanical loads and lesser BAT in females provided lesser osteogenic stimulus than greater BAT and lesser mechanical loads in males. It implies that at a constant body weight, the taller an individual is the greater the individual’s BMD. The vice versa is also true. Therefore, when prescribing skeletal muscle loading for improved bone health, individuals with varied heights should not be placed on the same biomechanical loading regimen.

We found no relationship between age and BMD (toe and thumb) in our study. The overall mean age for this study was 37.2±9.79 years suggesting that an average participant in this study was within the age group that is physiologically protected from experiencing significant bone loss. This is reasonable because the age-related bone mass increment peaks between ages 25 and 30 years [1], maintained for about 10 years [5, 86]. Thereafter the bone mass starts to decline [75] at about 0.3-0.5 % annually for both sexes [5,86] and rapidly in women between the ages of 45-55 years due to menopausal changes in estrogen production [5] with a consequent reduction in the BMD [75]. Therefore, in PLWH of similar age as participants in our study, the BMD value may not be affected by age.

We found no significant effect of the duration of HAART exposure on the BMD of PLWH. Of the 503 participants in our study, only a total of 352 participants were HAART-experienced with a mean exposure of 4.54±3.51 years suggesting that an average participant was on HAART for nearly 5 years. However, we found an important trend that could have some implications in a wider population study including that the highest cases of osteopenia (50% or 3 out of 6) occurred in those exposed to HAART for about 0 – 4 years, and 5 – 9 years, respectively. Similarly, the mean BMD values suggested that cases of osteopenia were common in non-weight bearing bones (i.e., the thumb unlike weight-bearing bones) of participants exposed to HARRT for 0 – 4 years (−1.04), 5 – 9 years (−1.07) and > 14 years (−1.22).

The only case of osteoporosis was reported among PLWH exposed to HAART for 5 – 9 years. Overall, only a small number (19 out of 475 or 4 %) of participants in our study presented with bone changes (osteopenia/osteoporosis) for exposure to HAART. This trend may agree with a previous finding by Cassetti et al. [34] of an increase of 1.6 % in BMD at the lumbar spine and a decrease of 0.1 % in the hip area between 48 weeks and 5.5 years of exposure to HAART. This is consistent with our findings that osteopenia occurred in 6/256 or 2.34 % of the participants between 0 – 4 years, while osteoporosis occurred in 1/256 or 0.4 % of the participants between 5 – 9 years, but no cases were recorded from 10 – 14 years or > 14 years. However, these values would have been slightly higher as only one of the seven participants who had osteoporosis provided information on the duration of exposure to HAART. A previous longitudinal study [26] found a decrease in BMD after 48 weeks and subsequently an increase in BMD after 96 weeks at the lumbar spine. In contrast, a meta-analysis study on stable bone density in HAART-treated HIV patients showed no consistent evidence of significant BMD loss for 1.5-2.7 years in studies involving PLWH [35]. Furthermore, Bolland et al., [35] also observed that commencing HAART at the onset of illness resulted in a stable or fairly increased BMD during the first year and stale or slightly decreased BMD in the second year. This stability is maintained till about two and half years before the onset of instability in BMD occurs [35] in HAART-experienced PLWH, unlike HAART-naïve PLWH. It suggests that early use of HAART contributes to the stability of the BMD in PLWH.

The classes of HAART (PI, NNRTI, and/or NRTI) seemed not to affect the BMD of PLWH. Thus, 267 (i.e., 75.9 %) participants out of 352 (who provided information on their drug history), were exposed to combination therapy of NRTI and NNRTI. In our study, we classified participants into four subgroups as regards the type of HAART taken, including NRTI, NNRTI, PI, as well as NRTI and NNRTI combination therapy. We found no association between the classes of HAART exposure and BMD irrespective of whether it was a weight-bearing bone or non-weight bearing bone. The heterogeneity of results in the literature [11, 25-30] may be due to the level of treatment adherence and also the type of HAART that the patients were exposed to (NNRTI, NRTI and/or PI). Importantly, it could be that the effects of the types of HAART on BMD are amplified or modulated in the presence of other risk factors, and should be interpreted in that context. Ordinarily, the most effective combination therapy used in suppressing the emergence of the resistant virus is a combination of three or more reverse transcriptase and protease inhibitors [87]. However, there is a general perception that HAART lowers BMD levels initially, subsequently stabilizes it and later on increases the BMD as a long-term effect [31]. A similar finding was reported by previous authors [87] such that the choice of antiretroviral (NRTI, NNRTI and/or PIs) did not have any effect or influence on BMD loss. Another study [32] found no significant association between BMD and combination antiretroviral therapy (CART) category (between protease inhibitor-based and non-nucleoside reverse transcriptase inhibitor-based). In another study [33] among HIV-positive women, 84 % had taken antiretrovirals, and 62% had taken protease inhibitors, but the class of HAART used was not associated with BMD. Another study [31] found that exposure to a tenofovir-based ART regimen was not significantly (p=0.417) associated with BMD. In contrast, a more recent study [4] found that tenofovir-based regimes were mostly associated with low BMD (52.4 %), and reported significant differences in the BMD in ART-naïve and ART-experienced patients. The authors concluded that since osteopenia and osteoporosis were highly prevalent in HAART-naïve patients, it seems that viremia if not controlled appropriately, might impact the BMD via its effects in inducing persistent systemic inflammation and bone remodelling.

## Conclusion

### Implication for practice

Our key findings suggest that weight-bearing bones (toes) were more protected from demineralization unlike the non-weight-bearing bones (thumb). It suggests that skeletal limb loading which occurs in walking or weight-bearing physical activities could be beneficial for bone health in HIV conditions. Our findings also suggest that if PLWH are at risk of developing an increased level of bone fragility, it could be a result of bone loss due to a multifactorial aetiology including sex differences, and body composition variations (specifically, BMI and height). Thus, we found that males were less likely to experience bone loss as much as females. This may warrant that PLWH who are females should be targeted as being at greater risk of bone loss or consequent fracture than males. The negative relationship between the underweight BMI category and BMD suggests that factors which alter the fat metabolism could also influence bone metabolism in HIV conditions. The significant effect of BMI on BMD was most widespread in PLWH who were normal-weight versus underweight and then followed by overweight versus underweight and obese versus underweight, in that order. Therefore, being underweight could be a physiological correlate of poor bone health and should be explored in future studies in PLWH.

Similarly, the findings of our study are of clinical importance because the literature reports that HIV has direct negative effects on bone mineral density as well as certain ARVs, including protease inhibitors, nucleoside/nucleotide reverse transcriptase inhibitors, and non-nucleoside reverse transcriptase inhibitors. [88-94]. These drugs were also used by the participants in our study from zero to 16 years duration (with an average duration of up to 4.54 years), yet we found that the type of HAART and duration of exposure to HAART have no significant negative relationship with the BMD. However, HAART usage from 5 – 9 years was associated with a case of osteoporosis. This could be an important indicator that HAART usage may create instability in the bone turnover from 5 – 9 years, and should be monitored in future studies. Considering that the average age of our study participants was 37 years, it is not clear whether a similar or more negative findings would be reported in older adults.

### Implications for future studies

The findings of our study highlight the potential of weight-bearing physical activities in enhancing skeletal muscle loading as a non-pharmacological prevention strategy against bone loss in PLWH. However, it is not known whether weight-bearing physical exercise activities could overcome the differences in BMD in PLWH attributed to low BMI (underweight). Therefore, high-quality randomized controlled trials (RCT) are required to provide an estimate of the effect of weight-bearing physical exercise activities on BMD in PLWH to guide practice. Also, the most effective exercise prescriptions that would prevent bone loss or improve BMD in PLWH should be determined in future high-quality RCTs. Presently, one RCT is ongoing in this area in Africa with sites in South Africa and Nigeria [58], while only one is completed [61] with a site in Malawi, but more is required, particularly in Africa which has the greatest global HIV burden [95].

### Strength and limitations

A key limitation of this study is that we recruited 503 participants that were available within the duration of the study which was lower than the calculated sample size of 760. The strength of our study is that it was conducted in a well-described population of PLWH who were HAART-experienced comprising males and females, young and old, placed on different types of HAART and with varied durations of HAART exposure ranging from zero to >15 years. Therefore, it was possible to stress relevant research questions that enabled answers that addressed the gaps in knowledge regarding the pattern of variations in BMD due to several traditional risk factors of interest. We also measured the BMD of weight-bearing and non-weight-bearing bones to elucidate the extent of BMD variations attributable to skeletal muscle loading and shear traction on the bones. This is the first time such a study is accomplished in Africa using an affordable technology - a portable Xrite 331C densitometer which is hand-held, can easily be deployed to clinics in remote villages and takes up to 2000 reads after charging the battery to full capacity. Therefore, it demonstrated the viability of using the densitometer in developing countries where power infrastructure is under-developed. The densitometer which has zero ionizing radiation [96] has been used in a previous study involving pre- and post-menopausal women [53], unlike the DEXA which has a low ionizing radiation exposure effect. The key limitation of our study is that it was a descriptive cross-sectional study and cannot establish a causal relationship between the study variables and BMD in PLWH. There were also incomplete data provided by participants which otherwise would have enabled a fuller understanding of the impact of the variables of interest on BMD in PLWH.

## Data Availability

The datasets generated and analyzed during the current study are not publicly available due to ethical concerns as the data contain potentially identifying and sensitive patient information but are available from the corresponding author and first author upon reasonable request.

## Acknowledgement

We wish to acknowledge with gratitude the management of Enugu State University Teaching Hospital, Parklane, Enugu, Nigeria, and staff of the HIV Clinic for all their support, permission, assistance, and contributions to the success of this work.

